# Trigger Point Injections for Post-Mastectomy Pain Syndrome: A Protocol for a Randomized Double-Blind Clinical Trial (NCT04267315)

**DOI:** 10.1101/2024.11.28.24317905

**Authors:** Victor Figueiredo Leite, Rodrigo Guimarães de Andrade, Christina May Moran de Brito

## Abstract

**INTRODUCTION:** Post-Mastectomy Pain Syndrome (PMPS) refers to a condition of chronic pain persisting for more than three months after a breast surgical procedure. It affects 11 to 70% of individuals with breast cancer. The pain has a mixed etiology, often with the frequent presence of associated myofascial pain. Trigger Point Injection (TPI) is a well-established procedure in the treatment of myofascial pain in the general population. However, there are no controlled studies evaluating the efficacy of TPI in the treatment of PMPS.

**OBJECTIVE:** To evaluate the efficacy of TPI in individuals with PMPS when combined with interdisciplinary rehabilitation and pharmacological treatment.

**METHODS:** This is the protocol for a double-blind, placebo-controlled clinical trial. Both groups will receive routine care by a Physiatrist and Rehabilitation Team blinded to the allocation. The active group will receive Trigger Point Injections with 1% lidocaine at each identified trigger point once a week for up to three consecutive weeks. The control group will receive subcutaneous infiltrations of saline at the same points and frequency. This protocol was registered at the ClinicalTrials.gov website (NCT04267315).

**STATISTICAL ANALYSIS:** Analysis of Covariance (ANCOVA) for between-group differences at baseline, one month, and three months for pain, central sensitization, and functionality (n=120). A significance level of alpha=5% and statistical power of 80% will be employed.

## 1. Introduction

Breast cancer is the most common neoplasm, excluding skin neoplasms ^(1)^. Individuals with breast cancer exhibit high rates of physical disabilities, with 60% experiencing symptoms such as pain or fatigue ^(2)^. Among the causes of pain, Post-Mastectomy Pain Syndrome (PMPS) is a significant entity ^(3)^.

PMPS is a condition of chronic pain occurring after a breast surgical procedure, persisting for more than three months post-surgery ^(3)^. It is a diagnosis of exclusion and does not encompass pain secondary to tumor recurrence or local infection, for example. PMPS is frequent, with an estimated incidence ranging from 11% to 70% among individuals undergoing breast cancer surgery ^(4-10)^. As a syndrome, PMPS has various presentations, with components of nociceptive, neuropathic, visceral pain, and central sensitization, varying in weight among different individuals ^(3, 11-18)^. Some individuals may present with a greater neuropathic pain component (e.g., incisional neuroma, intercostobrachial nerve neuropathy, phantom breast syndrome) ^(19, 20)^, while others have a greater nociceptive pain component (myofascial pain in the pectoral region, rotator cuff tendinopathy) ^(11-14)^.

Similarly to other chronic pain syndromes, the treatment of PMPS is multimodal, relying on interdisciplinary rehabilitation, pharmacological treatment, and invasive procedures ^(3, 21)^. In summary, the rehabilitation program aims to promote analgesia using physical modalities and kinesiotherapy; restore and improve range of motion (ROM); strengthen affected musculature; restore and enhance muscle coordination and functionality; and provide psychosocial support ^(3)^. Pharmacological treatment aims to reduce painful stimuli at various points and stimulate the descending analgesic system. Some evidence supports the efficacy of venlafaxine, gabapentin, amitriptyline, and topical capsaicin in PMPS treatment ^(3)^. Despite these treatment options, a considerable number of individuals continue to experience pain ^(21)^. The inclusion of Trigger Point Injection (TPI) in the multimodal treatment of PMPS may enhance treatment efficacy.

TPI is a cornerstone in myofascial pain treatment ^(22)^. Myofascial pain occurs when there is an imbalance between a specific muscle’s capacity and the applied load. Consequently, there is muscular contraction that compresses the intramuscular vascular-nerve bundle, creating a vicious cycle promoting increased muscle contraction. TPI induces immediate muscle relaxation, contributing to breaking this cycle of myofascial pain. Several studies indicate the presence of myofascial pain in individuals with breast cancer undergoing surgery ^(11-14, 23, 24)^.

The effect of myofascial pain treatment in individuals with breast cancer has been analyzed in previous studies ^(14, 25-29)^. Among these studies, only three included an invasive procedure as an intervention ^(26, 28, 29)^. Vas et al. and Shin et al. conducted non-controlled studies showing the potential benefit of TPI in PMPS. However, these studies, besides lacking control, did not involve interdisciplinary rehabilitation. De Groef et al. conducted a randomized double-blind clinical trial that demonstrated no benefit in injecting botulinum toxin type A to the pectoralis major muscle. However, this study included a control group receiving a non-inert intervention (intramuscular saline infiltration), which may have reduced the main intervention’s effect and involved the assessment of only one muscle.

Therefore, there is a need for a controlled double-blind clinical trial with an appropriate control group, coupled with an interdisciplinary rehabilitation program, to analyze the effect of TPI in individuals with PMPS.

### Primary Objective

Evaluate the efficacy of Trigger Point Injection (TPI) in participants with Post-Mastectomy Pain Syndrome (PMPS) when combined with interdisciplinary rehabilitation and pharmacological treatment, assessing pain, central sensitization, and functionality.

### Secondary Objective

Identify subgroups of individuals who derive greater benefit from TPI within the PMPS population.

## 2 METHODS

### 2.1 Study Type

This is a double-blind, randomized, controlled clinical trial comparing the effects of TPI + physical rehabilitation + pharmacological treatment (ACTIVE) with sham TPI + physical rehabilitation + pharmacological treatment (PLACEBO), with intention-to-treat analysis. This protocol was registered at the ClinicalTrials.gov database under NCT04267315.

### 2.2 Study Location

Physiatry Outpatient Clinic and Rehabilitation Center of the Cancer Institute of the State of São Paulo (ICESP), located in São Paulo, São Paulo, Brazil, and the Hospital de Amor, in Barretos, São Paulo, Brazil.

### 2.3 Participants or Inclusion/Exclusion Criteria

Inclusion Criteria:

- Female participants with breast cancer.
- Diagnosis of PMPS, according to Wisotzky et al^(3)^.
  ▪ Pain lasting > 3 months in the breast, lateral chest wall, or ipsilateral shoulder to surgery.
- Pain rated as Numeric Rating Scale (NRS, 0-10) ≥ 4.
- At least one active trigger point in the following muscles: pectoralis major, upper trapezius, serratus anterior, levator scapulae, latissimus dorsi, and infraspinatus.

Exclusion Criteria:

- Less than 3 months since last radiation therapy session.
- Allergy to lidocaine or other local anesthetics.
- Active infection over the injection area.
- Unavailability to come to the hospital to receive the intervention.

### 2.4 Intervention

At the initial assessment (T0), individuals will be screened based on the eligibility criteria. An informed consent will be obtained from all participants.

All participants will continue to receive routine care provided by the institution’s Physiatrist, who will be blinded to their allocation. Routine care includes orientation, home exercises, medication, referral to rehabilitation therapy (physical therapy, occupational therapy, among others). The research team will be responsible for administering TPIs, either active or placebo, and for collecting outcome data.

#### 2.4.1 Procedure

Participants will receive TPI once a week for up to three consecutive weeks. The applicator will prepare 6 mL of 1% lidocaine for the ACTIVE group or 6 mL of 0.9% sodium chloride for the PLACEBO group. The solution will be drawn into a 10 mL syringe attached to a 0.70 x 30 mm needle (22G x 1 ¼”). The participant will only enter the room after the solution has been prepared and all medication vials have been stored.

Before the injection, the presence of trigger points will assessed by physical examination, as well as the current pain intensity (NRS). If the patient has either NRS=0 or no active trigger points on that day, the intervention will not performed that week, and the participant will be asked to return in one week for the remaining sessions, if there are any.

During the intervention, the participant will be positioned in a lateral decubitus, with the side to be treated facing up, with their backs to the researcher. After the injection, the syringe will be immediately discarded to prevent the participant from identifying the administered volume. Only then the participant will be allowed to change their position.

##### ACTIVE

Participants in the ACTIVE group will receive TPI with 1 mL of 1% lidocaine in each of the muscles identified with active trigger points on the application day, as described above. The infiltration method will be standardized among applicators as an “in-and-out” technique: the needle will be advanced into the trigger point, followed by solution injection and needle removal. The classic fan-like needling technique will not be employed, and repetitive needling will not be performed. While a local twitch response is considered desirable, its presence or absence will not be documented. This technique was chosen to optimize blinding in the study. Trigger points were standardized according to those most frequently identified by Travell and Simons ^(30)^, and were defined as follows:

- Upper Trapezius: Midpoint between C7 and the acromion, in the anterior portion of the muscle. Needle inserted obliquely, from lateral to medial, cranially to caudally.
- Levator Scapulae: At the bifurcation between the levator scapulae and the upper trapezius, cranial to C7 and the acromion. Needle inserted from posterior to anterior (Figure 1)
- Infraspinatus: Divide the scapular spine into four equal parts, starting from its medial portion. At the division between the 1st and 2nd quarter, 2 cm below the scapular spine. Needle inserted from posterior to anterior (Figure 2).
- Latissimus Dorsi: In the posterior axillary region, where it is possible to pinch the muscle belly and separate it from the axillary wall. Needle inserted from lateral to medial.
- Serratus Anterior: In the mid-axillary line, at the most sensitive point between the 5th and 6th ribs. Needle inserted obliquely, from posterior to anterior, from lateral to medial, in the portion of the muscle overlying the rib.
- Pectoralis Major: In the clavicular portion of the muscle, 0.5 to 1.0 cm anterior to the mid-axillary line. Needle inserted obliquely, from lateral to medial, from anterior to posterior.

**Figure 1.**
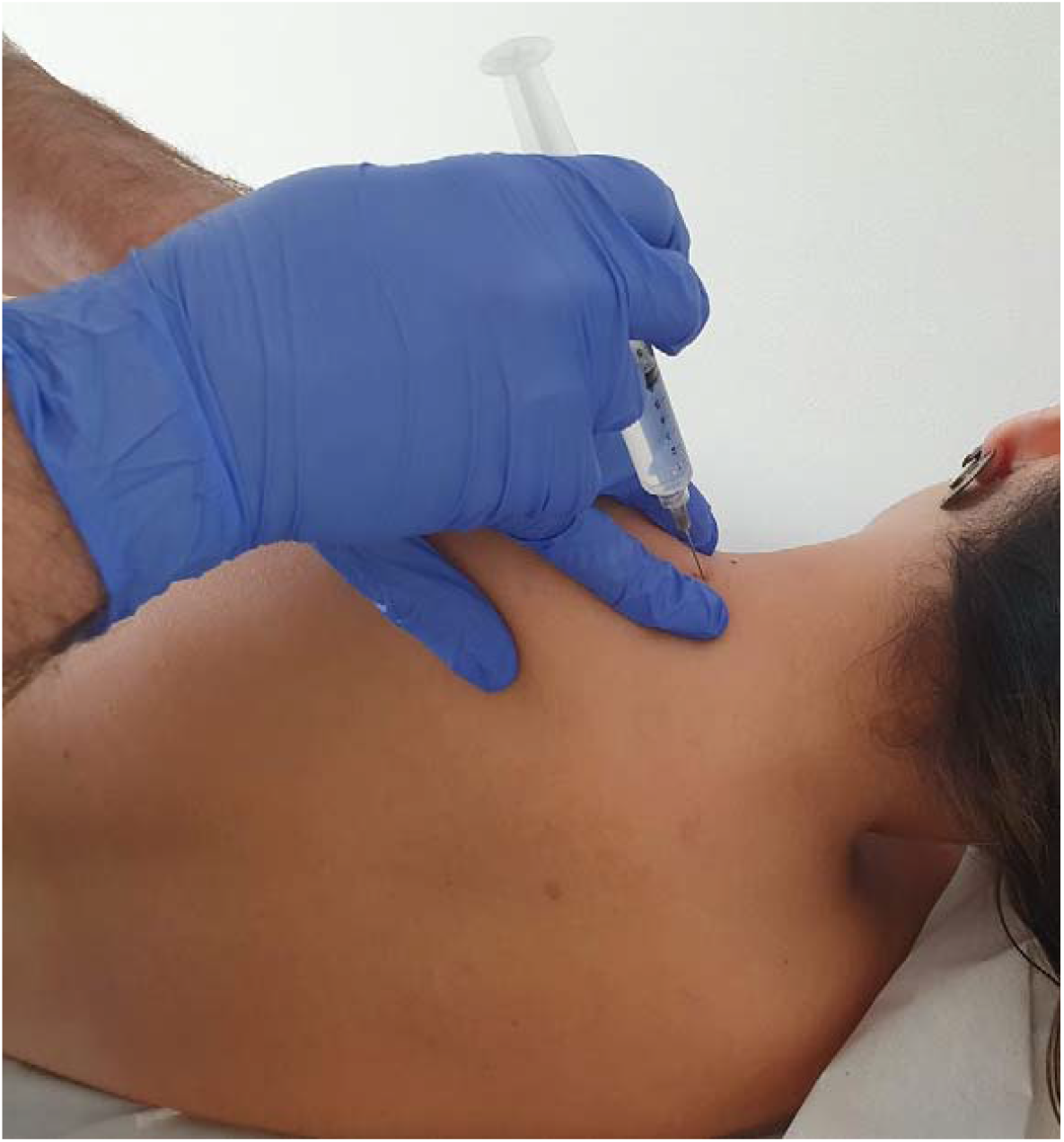
Levator scapulae injection technique

**Figure 2.**
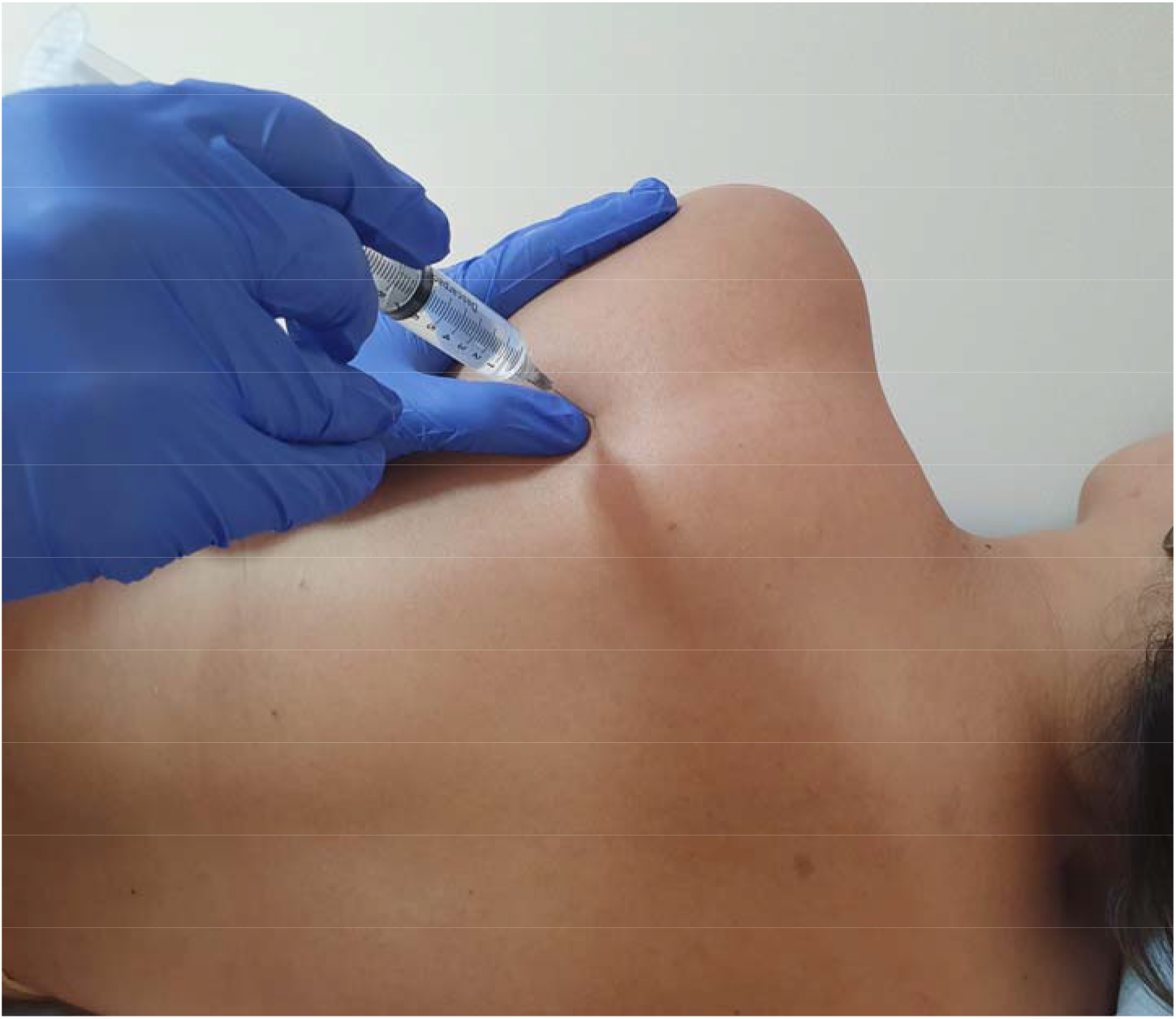
Infraspinatus injection technique

##### PLACEBO

Participants in the PLACEBO group will receive subcutaneous infiltration with 0.2 mL of 0.9% sodium chloride, administered superficially to each active trigger points on that specific session.

### 2.5 – Randomization, allocation and blinding

Participants will be randomized at the time of entry into the study, in permuted blocks of six, stratified by Axillary Clearance (binomial) and Reconstruction Surgery (binomial). The list will be created using an online software, “Sealed Envelope” ^(31)^, and only one of the authors (CMMB) will have access to the list until the completion of the study.

Before the first TPI session, the researcher performing the intervention will obtain the participant’s allocation from one of the authors (CMMB) via email or other electronic method. The evaluator, the participant, the institution’s Physiatrist, and the Rehabilitation team will not have access to the participant’s allocation.

The outcome assessor, the participant, the institutional Physiatrist, and the Rehabilitation team will remain blinded to participant allocation until the end of the study. Only the researcher who performed the infiltration and the supervising researcher will have access to this information.

### 2.6 Outcomes

All questionnaires have been translated and validated to Brazilian Portuguese.

#### 2.6.1 Primary outcomes

Difference in Pain between baseline and 3 months (T3m) for both ACTIVE and PLACEBO groups, measured by:

- Numeric rating scale (NRS, 0-10) for average pain in the last few days.
- Pressure pain threshold over the six muscles previously described, both ipsilateral and contralateral to side being treated, assessed in kg/cm^2^.
  ∘ Evaluated in the physical examination using a validated digital pressure algometer (Med.Dor, Gonçalves, Minas Gerais, Brazil) ^(32)^.
  ∘ Pressure applied to the trigger point perpendicular to the muscle, starting at 0 kg/cm2 up to a maximum of 5 kg/cm2. Three tests were performed on each muscle, bilaterally, with at least 30-second interval between each measurement in the same muscle ^(33, 34)^. The average value of the three measurements was used for analysis.

Incidence of active trigger points

- Assessed by palpation during physical exam, as per Simons and Travell criteria ^(30)^.

##### Upper trapezius

- Midpoint between C7 apophysis, and acromium, at the anterior portion of the muscle, pressed from cranial to caudal (Figure 3).

**Figure 3.**
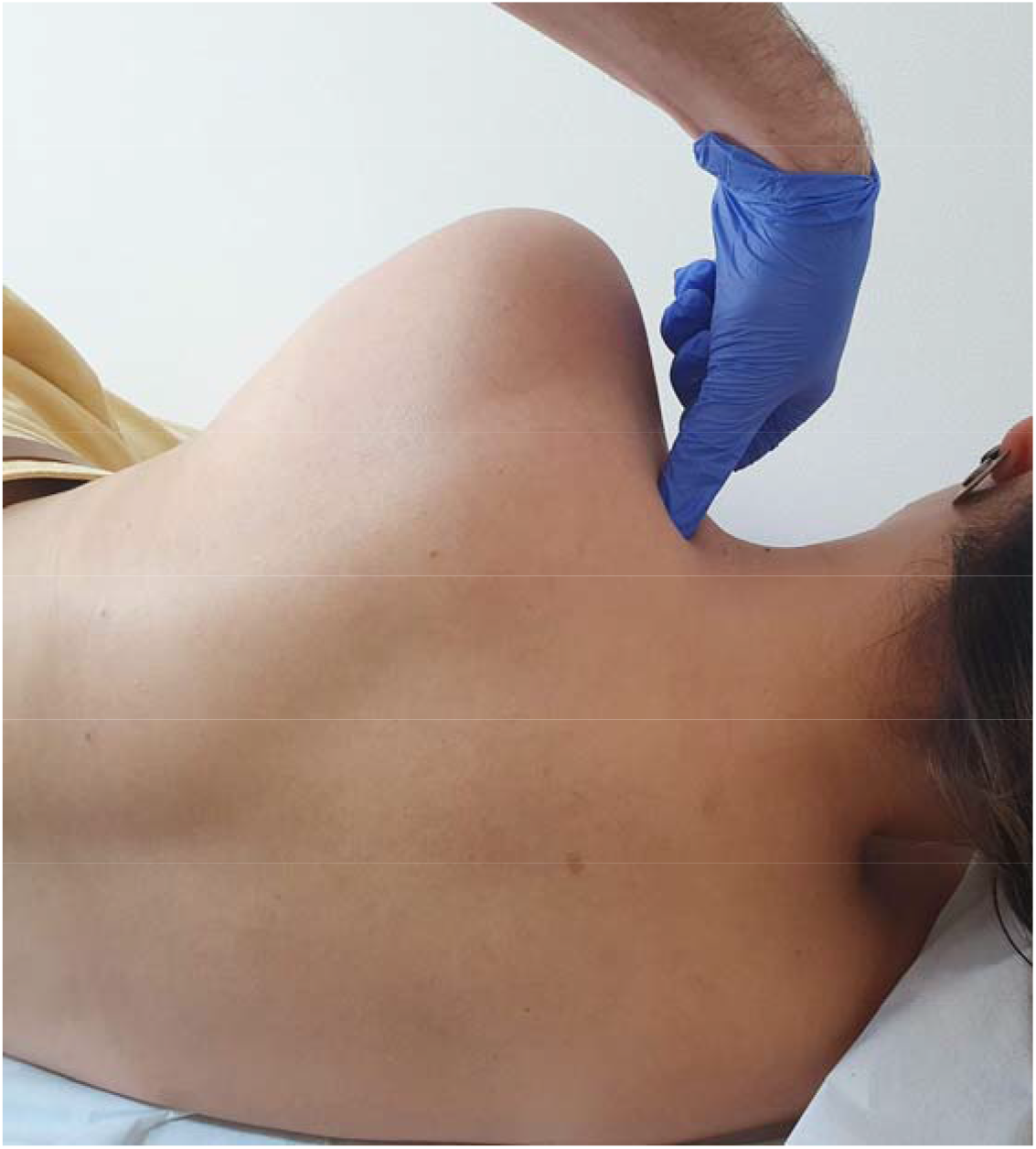
Upper trapezius trigger-point assessment

##### Levator scapulae

- In the bifurcation between levator scapulae and upper trapezius, cranial to the line between C7 and the acromium. Pressed from lateral to medial.

##### Infraespinatus

- Divide the scapular spine into four equal portions, starting from its medial part. At the division between the 1st and 2nd quarters, 2cm below the scapular spine. Apply pressure from posterior to anterior.

##### Latissimus Dorsi

- In the posterior axillary region, where its muscular belly can be pinched and separated from the axillary wall. Apply pressure from lateral to medial.

##### Serratus Anterior

- In the mid-axillary line, at the most sensitive point between the 5th and 6th ribs. Apply pressure from lateral to medial, between the ribs.

##### Pectoralis Major

- In the clavicular portion of the muscle, 0.5 to 1.0cm anterior to the anterior axillary line. Apply pressure from anterior to posterior while holding the muscular belly with the other hand.

#### 2.6.2 Secondary outcomes

Difference between baseline (T0) and 1 month (T1m), and T0 and T3m for:

- NRS: absolute difference, 30% and 50% improvement.
- Short-Form McGill Pain Questionnaire (SF-MPQ) – Total score, and in sensory and affective categories.
- Neuropathic Pain Symptom Inventory (NPSI) ^(35)^.
- Disabilities of the Arm, Shoulder and Hand Questionnaire (DASH) ^(36)^.
- Central Sensitization Inventory (CSI) ^(37, 38)^.
- Range of motion (ROM) for shoulder for abduction and external rotation.
  ∘ Ipsilateral to the treated site
  ∘ Participant will be positioned sitting in a chair or examination table. External rotation will be examined with the shoulder neutral to abduction.
- Self-reported global perception of improvement (Likert scale, 1 to 7)
- Adverse events
- Use of pain medication
  ∘ Subjectively classified as “Increase,” “Stable,” “Reduction,” or “Unable to determine.”

### 2.7 Statistical analysis

Continuous variables will be described as mean and standard deviation (SD); ordinal variables as median and interquartile range; and categorical variables as the percentage of incidence. Continuous variables will be assessed using the Kolmogorov-Smirnov normality test.

The primary analysis of the study will be performed by Analysis of Covariance (ANCOVA). The outcome will be the difference in NRS between groups (ACTIVE-PLACEBO) between T0 and T3m; with the following covariates in the model: NRS at T0, age, BMI, lymphedema, radiotherapy, chemotherapy, type of surgery, breast reconstruction, smoking, central sensitization (defined as CSI>40), HADS, and axillary clearance (binomial).

The secondary analyses will be performed using ANCOVA with identical covariates to the primary, for both T1 and T3, for the following outcomes:

- NRS
- NRS30%
- NRS50%
- SF-MPQ (total, sensory, and affective)
- NPSI
- DASH
- CSI
- ROM
- GPE
- Adverse events
- Use of pain medication

Participants will be assessed using an intention-to-treat analysis. In case of loss of follow-up, data imputation methods will be used.

#### 2.7.1 – Sample Size Calculation

The expected effect size of the intervention between T3m and T0 is -2.0, which is the minimum clinically important difference in this population using this scale ^(39)^. The SD was obtained using data from several studies in the PMPS population ^(22, 25-27)^. From this range between 5.5 to 8.4, we chose the value of

The following parameters were used:

- Effect size = 2.0
- SD = 7.1
- Alpha = 5%
- Statistical power = 80%
- Partial sample size = 101

Considering a 20% participant loss, we determined:

- Total sample size = 120

## 3 - Discussion

This is the protocol of a double-blind, placebo-controlled clinical trial investigating the efficacy of TPI in the treatment of PMPS when combined with interdisciplinary rehabilitation and pharmacological treatment. PMPS, characterized by a complex interplay of nociceptive, neuropathic, and myofascial pain components, often proves challenging to manage with pharmacological treatments and physical rehabilitation alone ^(3, 16, 21)^.

Comparing this study’s approach to existing literature reveals a nuanced understanding of PMPS treatment. Previous studies have explored various interventions, ranging from pharmacological approaches to physical therapies, with mixed results. The inclusion of a placebo group and the focus on a combination of TPI with rehabilitation and pharmacology differentiate this study, offering new insights into how combining treatments can enhance patient outcomes. This trial thus will contribute to a growing body of evidence suggesting that no single modality may suffice to address the multifaceted nature of PMPS pain, advocating for a more holistic, patient-centered approach to management.

## Data Availability

All data produced in the present study will be available upon reasonable request to the authors

## Notes

### Competing Interest Statement

The authors have declared no competing interest.

### Clinical Trial

NCT04267315

### Funding Statement

This study did not receive any funding

## References

1. Instituto Nacional de Câncer José Alencar Gomes da Silva (INCA). Estimativa 2023: incidência de câncer no Brasil. In: Vigilância. CdPe, editor. Rio de Janeiro: INCA; 2022.

2. Cheville AL, Mustian K, Winters-Stone K, Zucker DS, Gamble GL, Alfano CM. Cancer Rehabilitation: An Overview of Current Need, Delivery Models, and Levels of Care. Phys Med Rehabil Clin N Am. 2017;28(1):1–17.

3. Wisotzky E, Hanrahan N, Lione TP, Maltser S. Deconstructing Postmastectomy Syndrome: Implications for Physiatric Management. Phys Med Rehabil Clin N Am. 2017;28(1):153–69.

4. Mejdahl MK, Andersen KG, Gartner R, Kroman N, Kehlet H. Persistent pain and sensory disturbances after treatment for breast cancer: six year nationwide follow-up study. BMJ. 2013;346:f1865.

5. Romero A, Tora-Rocamora I, Bare M, Barata T, Domingo L, Ferrer J, et al. Prevalence of persistent pain after breast cancer treatment by detection mode among participants in population-based screening programs. BMC cancer. 2016;16(1):735.

6. Gartner R, Jensen MB, Nielsen J, Ewertz M, Kroman N, Kehlet H. Prevalence of and factors associated with persistent pain following breast cancer surgery. JAMA. 2009;302(18):1985–92.

7. Vilholm OJ, Cold S, Rasmussen L, Sindrup SH. The postmastectomy pain syndrome: an epidemiological study on the prevalence of chronic pain after surgery for breast cancer. Br J Cancer. 2008;99(4):604–10.

8. Belfer I, Schreiber KL, Shaffer JR, Shnol H, Blaney K, Morando A, et al. Persistent postmastectomy pain in breast cancer survivors: analysis of clinical, demographic, and psychosocial factors. J Pain. 2013;14(10):1185–95.

9. Juhl AA, Christiansen P, Damsgaard TE. Persistent Pain after Breast Cancer Treatment: A Questionnaire-Based Study on the Prevalence, Associated Treatment Variables, and Pain Type. J Breast Cancer. 2016;19(4):447–54.

10. Edmond SN, Shelby RA, Keefe FJ, Fisher HM, Schmidt JE, Soo MS, et al. Persistent Breast Pain Among Women With Histories of Breast-conserving Surgery for Breast Cancer Compared With Women Without Histories of Breast Surgery or Cancer. Clin J Pain. 2017;33(1):51–6.

11. Torres Lacomba M, Mayoral del Moral O, Coperias Zazo JL, Gerwin RD, Goni AZ. Incidence of myofascial pain syndrome in breast cancer surgery: a prospective study. Clin J Pain. 2010;26(4):320–5.

12. Fernandez-Lao C, Cantarero-Villanueva I, Fernandez-de-Las-Penas C, Del-Moral-Avila R, Menjon-Beltran S, Arroyo-Morales M. Development of active myofascial trigger points in neck and shoulder musculature is similar after lumpectomy or mastectomy surgery for breast cancer. J Bodyw Mov Ther. 2012;16(2):183–90.

13. Caro-Moran E, Fernandez-Lao C, Diaz-Rodriguez L, Cantarero-Villanueva I, Madeleine P, Arroyo-Morales M. Pressure Pain Sensitivity Maps of the Neck-Shoulder Region in Breast Cancer Survivors. Pain Med. 2016;17(10):1942–52.

14. Castro-Martin E, Ortiz-Comino L, Gallart-Aragon T, Esteban-Moreno B, Arroyo-Morales M, Galiano-Castillo N. Myofascial Induction Effects on Neck-Shoulder Pain in Breast Cancer Survivors: Randomized, Single-Blind, Placebo-Controlled Crossover Design. Arch Phys Med Rehabil. 2017;98(5):832–40.

15. De Groef A, Meeus M, De Vrieze T, Vos L, Van Kampen M, Geraerts I, et al. Unraveling Self-Reported Signs of Central Sensitization in Breast Cancer Survivors with Upper Limb Pain: Prevalence Rate and Contributing Factors. Pain Physician. 2018;21(3):E247–e56.

16. Leysen L, Adriaenssens N, Nijs J, Pas R. Chronic pain in breast cancer survivors: nociceptive, neuropathic or central sensitization pain? 2018.

17. Galiano-Castillo N, Fernandez-Lao C, Cantarero-Villanueva I, Fernandez-de-Las-Penas C, Menjon-Beltran S, Arroyo-Morales M. Altered pattern of cervical muscle activation during performance of a functional upper limb task in breast cancer survivors. Am J Phys Med Rehabil. 2011;90(5):349–55.

18. Shamley D, Lascurain-Aguirrebena I, Oskrochi R. Clinical anatomy of the shoulder after treatment for breast cancer. Clin Anat. 2014;27(3):467–77.

19. Bouhassira D, Luporsi E, Krakowski I. Prevalence and incidence of chronic pain with or without neuropathic characteristics in patients with cancer. Pain. 2017;158(6):1118–25.

20. Mulvey MR, Boland EG, Bouhassira D, Freynhagen R, Hardy J, Hjermstad MJ, et al. Neuropathic pain in cancer: systematic review, performance of screening tools and analysis of symptom profiles. Br J Anaesth. 2017;119(4):765–74.

21. Tait RC, Zoberi K, Ferguson M, Levenhagen K, Luebbert RA, Rowland K, et al. Persistent Post-Mastectomy Pain: Risk Factors and Current Approaches to Treatment. J Pain. 2018.

22. Liu L, Huang QM, Liu QG, Ye G, Bo CZ, Chen MJ, et al. Effectiveness of dry needling for myofascial trigger points associated with neck and shoulder pain: a systematic review and meta-analysis. Arch Phys Med Rehabil. 2015;96(5):944–55.

23. De Groef A, Van Kampen M, Dieltjens E, De Geyter S, Vos L, De Vrieze T, et al. Identification of Myofascial Trigger Points in Breast Cancer Survivors with Upper Limb Pain: Interrater Reliability. Pain Med. 2018;19(8):1650–6.

24. Cantarero-Villanueva I, Fernandez-Lao C, Fernandez-de-Las-Penas C, Lopez-Barajas IB, Del-Moral-Avila R, de la-Llave-Rincon AI, et al. Effectiveness of water physical therapy on pain, pressure pain sensitivity, and myofascial trigger points in breast cancer survivors: a randomized, controlled clinical trial. Pain Med. 2012;13(11):1509–19.

25. De Groef A, Van Kampen M, Vervloesem N, Dieltjens E, Christiaens MR, Neven P, et al. Effect of myofascial techniques for treatment of persistent arm pain after breast cancer treatment: randomized controlled trial. Clin Rehabil. 2018;32(4):451–61.

26. De Groef A, Devoogdt N, Van Kampen M, Nevelsteen I, Smeets A, Neven P, et al. Effectiveness of Botulinum Toxin A for Persistent Upper Limb Pain After Breast Cancer Treatment: A Double-Blinded Randomized Controlled Trial. Arch Phys Med Rehabil. 2018;99(7):1342–51.

27. Rangon FB, Koga Ferreira VT, Rezende MS, Apolinario A, Ferro AP, Guirro ECO. Ischemic compression and kinesiotherapy on chronic myofascial pain in breast cancer survivors. J Bodyw Mov Ther. 2018;22(1):69–75.

28. Shin HJ, Shin JC, Kim WS, Chang WH, Lee SC. Application of ultrasound-guided trigger point injection for myofascial trigger points in the subscapularis and pectoralis muscles to post-mastectomy patients: a pilot study. Yonsei Med J. 2014;55(3):792–9.

29. Vas L, Pai R. Ultrasound-Guided Dry Needling As a Treatment For Postmastectomy Pain Syndrome - A Case Series of Twenty Patients. Indian journal of palliative care. 2019;25(1):93–102.

30. Simons DG, Travell JG, Simons LS. Travell & Simons’ myofascial pain and dysfunction : the trigger point manual. 2nd ed ed. Baltimore: Williams & Wilkins; 1999.

31. Sealed Envelope Ltd. 2017. Create a blocked randomisation list. [Online] Available from: https://www.sealedenvelope.com/simple-randomiser/v1/lists x[Accessed 30 Oct 2018].

32. Jerez-Mayorga D, Dos Anjos CF, Macedo MC, Fernandes IG, Aedo-Muñoz E, Intelangelo L, et al. Instrumental validity and intra/inter-rater reliability of a novel low-cost digital pressure algometer. PeerJ. 2020;8:e10162.

33. Walton DM, Levesque L, Payne M, Schick J. Clinical pressure pain threshold testing in neck pain: comparing protocols, responsiveness, and association with psychological variables. Physical therapy. 2014;94(6):827–37.

34. Bisset LM, Evans K, Tuttle N. Reliability of 2 protocols for assessing pressure pain threshold in healthy young adults. Journal of manipulative and physiological therapeutics. 2015;38(4):282–7.

35. de Andrade DC, Ferreira KA, Nishimura CM, Yeng LT, Batista AF, de Sa K, et al. Psychometric validation of the Portuguese version of the Neuropathic Pain Symptoms Inventory. Health Qual Life Outcomes. 2011;9:107.

36. Orfale A.G. APMP, Ferraz M.B., Natour J. Translation into Brazilian Portuguese, cultural adaptation and evaluation of the reliability of the Disabilities of the Arm, Shoulder and Hand Questionnaire. . Braz J Med Biol Res 2005;38(2):293–302.

37. Caumo W, Antunes LC, Elkfury JL, Herbstrith EG, Busanello Sipmann R, Souza A, et al. The Central Sensitization Inventory validated and adapted for a Brazilian population: psychometric properties and its relationship with brain-derived neurotrophic factor. J Pain Res. 2017;10:2109–22.

38. Scerbo T, Colasurdo J, Dunn S, Unger J, Nijs J, Cook C. Measurement Properties of the Central Sensitization Inventory: A Systematic Review. Pain Pract. 2018;18(4):544–54.

39. Harrington S, Gilchrist L, Sander A. Breast Cancer EDGE Task Force Outcomes: Clinical Measures of Pain. Rehabilitation oncology (American Physical Therapy Association Oncology Section). 2014;32(1):13–21.

